# Evidence for probable aerosol transmission of SARS-CoV-2 in a poorly ventilated restaurant

**DOI:** 10.1101/2020.04.16.20067728

**Authors:** Yuguo Li, Hua Qian, Jian Hang, Xuguang Chen, Ling Hong, Peng Liang, Jiansen Li, Shenglan Xiao, Jianjian Wei, Li Liu, Min Kang

## Abstract

**Background:** The role of aerosols in the transmission of SARS-CoV-2 remains debated. We analysed an outbreak involving three non-associated families in Restaurant X in Guangzhou, China, and assessed the possibility of aerosol transmission of SARS-CoV-2 and characterize the associated environmental conditions.

**Methods:** We collected epidemiological data, obtained a video record and a patron seating-arrangement from the restaurant, and measured the dispersion of a warm tracer gas as a surrogate for exhaled droplets from the suspected index patient. Computer simulations were performed to simulate the spread of fine exhaled droplets. We compared the in-room location of subsequently infected cases and spread of the simulated virus-laden aerosol tracer. The ventilation rate was measured using the tracer decay method.

**Results:** Three families (A, B, C), 10 members of which were subsequently found to have been infected with SARS-CoV-2 at this time, or previously, ate lunch at Restaurant X on Chinese New Year’s Eve (January 24, 2020) at three neighboring tables. Subsequently, three members of family B and two members of family C became infected with SARS-CoV-2, whereas none of the waiters or 68 patrons at the remaining 15 tables became infected. During this occasion, the ventilation rate was 0.75–1.04 L/s per person. No close contact or fomite contact was observed, aside from back-to-back sitting by some patrons. Our results show that the infection distribution is consistent with a spread pattern representative of exhaled virus-laden aerosols.

**Conclusions:** Aerosol transmission of SARS-CoV-2 due to poor ventilation may explain the community spread of COVID-19.

## Introduction

Debate continues on the role of aerosol transmission of SARS-CoV-2, the virus that causes COVID-19, in the rapidly growing COVID-19 pandemic. The COVID-19 outbreak in Guangzhou, China was identified in early 2020 and linked to three seemingly non-associated clusters of unrelated families (A, B, C) (Lu et al., 2020). Families B (*n* = 4) and C (*n* = 7) comprised local Guangzhou residents with no history of travel to or encounters with inhabitants from Hubei, but nevertheless three members of family B and two members of family C were confirmed infected with the virus on February 4 or 6, at which time only ∼10 cases of infection had been confirmed in the city.

Local health officials learned that families B and C had eaten lunch at the same restaurant on Chinese New Year’s Eve (January 24, 2020), as had family A (*n* = 10) from Wuchang, Wuhan (the epicenter of the Chinese epidemic), who had arrived in Guangdong by train on January 23. One person from family A reported experiencing the onset of COVID-19 symptoms on January 24, and video records from the restaurant show that families A, B, and C were seated at tables along the exterior window, with family A’s table in the center. None of the restaurant waiters or remaining 68 patrons distributed at 15 other tables became infected with SARS-CoV-2. Families A, B, and C had not met previously and did not have close contact during the lunch, aside from some patrons sitting back-to-back.

To investigate the possibility of aerosol transmission of SARS-CoV-2, we analyzed the spatial distribution data from this outbreak using computer models and experiments based on airflow dynamics. We use our results to assess the ventilation conditions of aerosol transmission.

## Methods

### Epidemiologic analysis

We obtained the seating arrangement of the three family members and remaining patrons in Restaurant X as well as the dates of COVID-19 symptom onset (Figure 2A), where the symptom onset date is defined as the day when symptoms (e.g., fever or cough) were first noticed by the patient. SARS-CoV-2 infection was confirmed by real-time polymerase chain reaction with reverse transcription (RT-PCR) analysis of throat swabs. Demographic data, travel history, exposure history, and symptoms of the infected individuals were collected (Lu et al., 2020), and we also obtained the floor plan and design of the air conditioning and ventilation system of the restaurant, and the hourly weather data for January 24 from a weather station near the site. A closed-circuit television camera record in the restaurant and elevator was reviewed to determine the elevator use by patrons, the fire-door use by both patrons and waiters, the table and seating arrangement during the lunch, and any close contact behavior between Family A and others.

**Figure 1.**
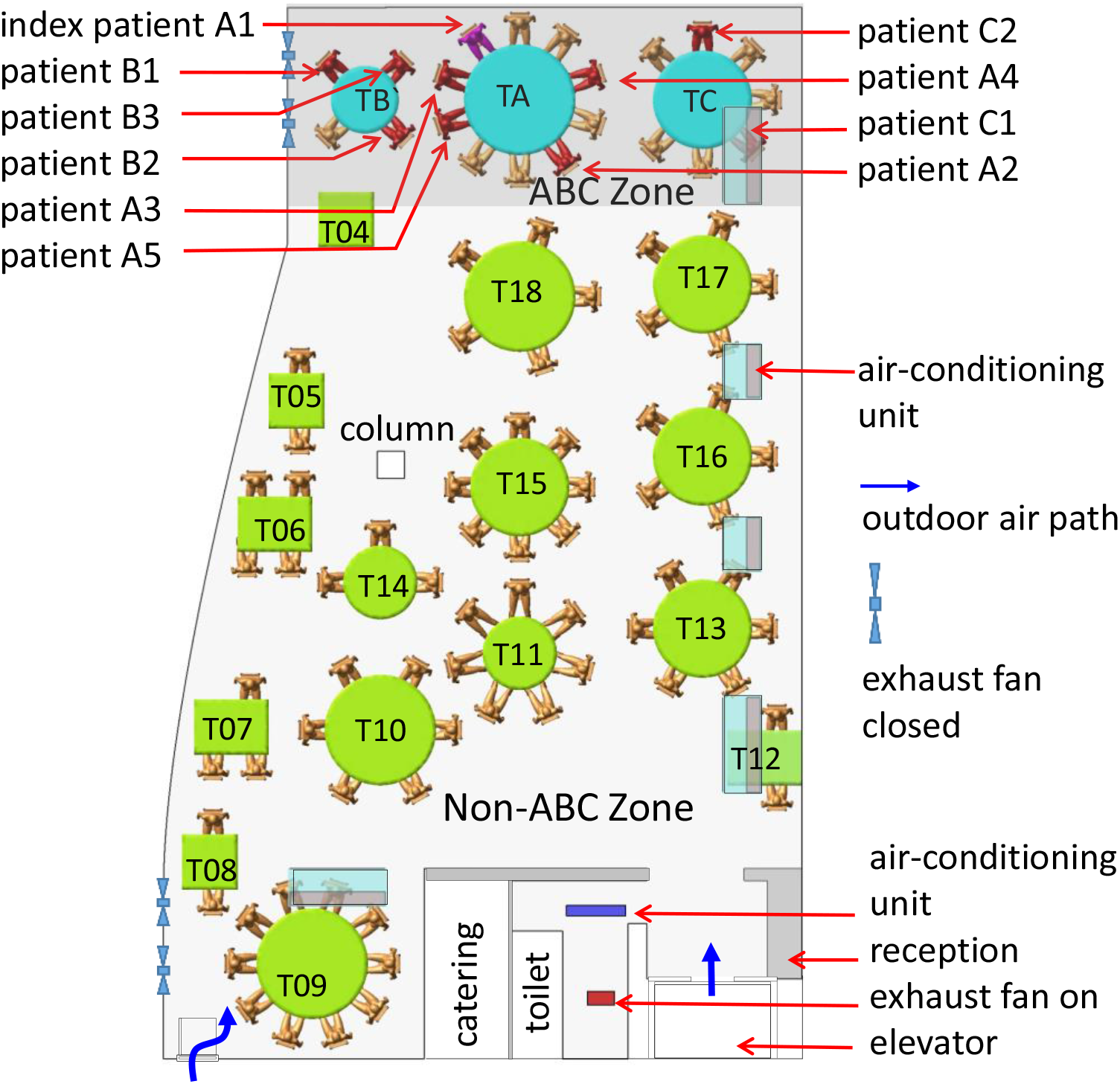
Distribution of SARS-CoV-2 infection cases at tables in Restaurant X. The probable air-flow zones are in dark grey and light grey. Each table is numbered as T#. Eighty-nine patrons are shown at the 18 tables, with one table being empty (T04). Tables TA, TB, and TC are where families A, B and C sat, some of whose members became infected. Patient A1 at TA is the suspected index patient. Patients A2–A5, B1–B3, and C1–C2 are the individuals who became infected. Other tables are numbered as T4–T18. Each of the five air-conditioning units condition a particular zone. Patrons and waiters entered the restaurant floor via the elevator and stairwell, which are connected by the fire door.

**Figure 2.**
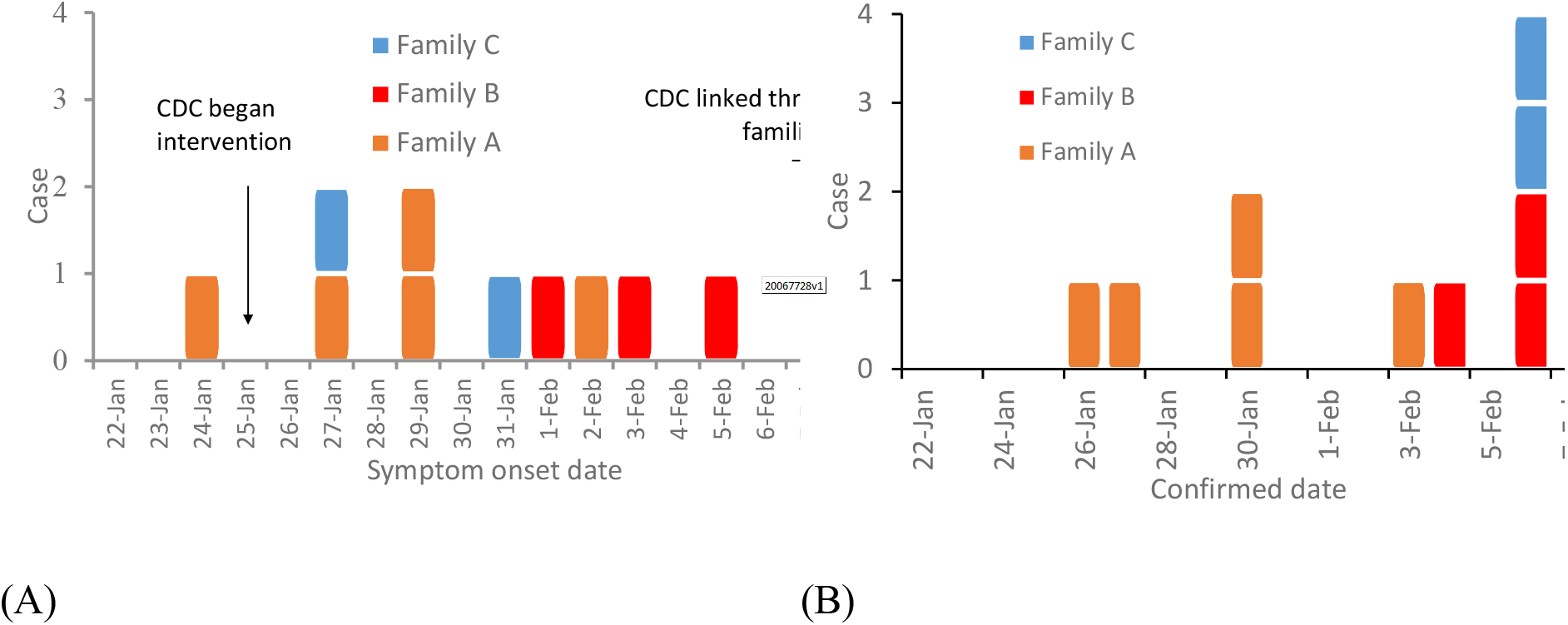
Dates of (A) symptom onset and (B) confirmation of the 10 patients from the three families.

Restaurant X has five floors. The outbreak occurred on the third floor, which has a volume of 431 m^3^ (height of 3.14 m, length of 17 m, and average width of 8.1 m) (Figure 1). Large and small tables have a diameter of 1.8 m and 1.2 m, respectively, and rectangular tables measure 0.9 m × 0.9 m and 1.2 m × 0.9 m. Five fan coil air-conditioning units are installed on the third floor, and there is no outdoor air supply: ventilation is thus achieved using only infiltration and natural ventilation through an occasionally open door driven by an exhaust fan installed inside the restroom. Four exhaust fans are installed on the south glass window but were not used during this lunch. At noon on January 24, the third floor of the restaurant had 18 tables and 89 patrons. We label Tables A, B, and C as TA, TB, and TC, respectively, and the remaining tables are labeled as T4–T18 (Figure 1). According to video analyses, the fire door was used approximately every 2 minutes.

We studied the infection data with regards to seating location and used a chi-square test to explore the relationship between a patron’s location (i.e., table) and his/her probability of becoming infected. Table A was excluded in this analysis. The other tables were categorized according to two criteria: distance from TA (e.g., immediate vs. remote neighbors) and air-conditioning zone (i.e., the ABC zone was that immediately around TA, TB and TC and serviced by one air conditioning unit, and the non-ABC zone was everywhere else, serviced by the four other air conditioning units).

### Experimental tracer gas measurements and computational fluid-dynamics simulations

Tracer gas measurements and computational fluid dynamics (CFD) simulations were used to predict the spread of fine droplets exhaled by the index patient and the detailed airflow pattern in the restaurant. The CFD simulation models were the same as those used in previous studies of two 2003 SARS-CoV outbreaks in Hong Kong (Yu et al., 2004, Wong et al., 2004, Li et al., 2005).

The tracer measurement was carried out on March 19–20 when the intensity of the direct solar radiation was similar to that on January 24, i.e., weak sunshine, with clouds and rain. We first measured the supply/return/exhaust air flow and temperature at each air-conditioning unit and at the exhaust fan in the restroom. We arranged the tables and chairs to match the arrangement used at the January 24 lunch, as determined by analyses of the video of this occasion. The air conditioning units were turned on and the exhaust fans in the vertical glass window were left off to simulate the air-flow conditions at the time of SARS-CoV-2 infection during the lunch on January 24. Volunteers were not recruited because the experiment was performed during the strict intervention (i.e., lockdown) phase of the epidemic in Guangzhou. However, nine experimental staff sat on Tables A, B, and C and simple thermal mannequins were placed at the others. The mannequins were warm and hollow, containing a 60-W electrical bulb enclosed by a stainless steel cylinder, which produced warm plumes similar to the above human subjects. The same simple heat source with a 60-W electrical bulb was also used to simulate warm food on each table.

The tracer gas measurement consisted of two stages. In the first stage, we released ethane gas through an 8-mm inner diameter pipe at a speed of 1.5 m/s at 32–34 °C, with the pipe outlet placed immediately above the index patient’s nose. This mode of release mimicked the index patient (assumed to be A1) talking and moving their head around. Tracer gas is known to be an effective surrogate for modelling the spread of fine droplets or droplet nuclei (Bivolarova et al., 2017). In the first of two experiments, we monitored the gas concentrations at 14 points, namely all of the chairs where the infected persons of families B and C had sat (Figure S3). In the second experiment, gas concentrations were only monitored at seven points, owing to the time required for rotational sampling at each point.

In the second stage, the air change rate was measured using the tracer decay method, which involved the release of a tracer gas into the restaurant that subsequently mixed the flow from with 10 desk fans. We measured the tracer concentration at three points in the room. The elevator and fire door were used every two minutes to mimic the traffic that was observed in the recording of the January 24 lunch, with the fire door closing automatically after a period of 3 s. We identified an exhaust flow through the doorway of the restroom and that bidirectional air exchange occurred through the opening and closing of the fire door. The non-operating exhaust fans were sealed relatively well, with nearly negligible air flow. After the measurements, we assigned the health status (ill vs. healthy) of each person at non-A tables as the dependent variable and applied a binary logistic regression model to investigate the association between the measured concentrations of trace gas and infection probability.

We adopted the widely used CFD software package Fluent (Ansys Fluent, USA), which is a three-dimensional, general-purpose CFD software package for modeling fluid flows. We used the basic renormalization-group (RNG) *k*-*ε* turbulence model to simulate the effects of turbulence on airflow and dispersion of pollutants. We assumed that the virus-laden water droplets generated from the index patient at TA rapidly evaporated (i.e., after a few s in air). We approximated the exhaled droplet nuclei as a passive scalar and the deposition effect was therefore neglected. The prediction was compared to measurement (Figure S4). After CFD modeling, we used the health status (ill vs. healthy) of each person at non-A tables as the dependent variable and applied a binary logistic regression model to investigate the association between the predicted concentrations and infection probability. In both the experiments and simulations, we assumed that the tracer gas was released from the index patient’s mouth.

## Results

### The outbreak

Detailed epidemiological, clinical, laboratory, and genomic findings for this outbreak and all of the associated patients have been described in detail by Lu et al. (2020). The first confirmed case (A1) from family A who was confirmed on January 24 is assumed to be the index patient. The last patient was confirmed on February 6 (Figure 2B). The three families occupied the restaurants at different times (family A 12:01–13:23; family B 11:37–12:54; and family C 12:03–13:18). According to the video analysis, there was no significant close contact between the three families in the elevator or restroom (Supplementary information A). Contact tracing identified 193 patrons in the restaurant, 68 of whom were on the third floor at the same time as families A, B, and C, including 57 restaurant workers and 11 workers in the hotel where Family A had stayed. None of these people were infected with the virus. Thus, only the 10 patrons in the restaurant were infected, comprising the index patient and nine others, and at least five of them who we suspect became infected at this lunch due to exposure to exhaled droplets from the index patient that contained virus particles.

### Spatial distribution analysis of infection cases

The tables and patrons were first categorized by distance from TA, as immediate neighbors (TB, TC, and T18) or remote neighbors (Tables T4–17). The 10 patients who were shortly thereafter be confirmed as having COVID-19 sat at one of the three tables by the window. Three of the four members of family B were infected, and two of the seven members of family C were infected. Five members of family A were also infected, including the index patient. The two patrons at TC who sat the closest to TA were not infected, nor were any patrons at the remote neighboring tables, but the patrons at neighboring tables had a higher infection probability than patrons at remote tables (*X*^*2*^*=*16.08, *P*<0.001, chi-squared test with continuity correction, Table 1). The infection risk was also higher for patrons at zone ABC tables than those at non-ABC zone tables (*X*^*2*^*=*25.78, *P*<0.001, chi-squared test with continuity correction, Table 1). None of the patrons seated in the non-ABC zone were infected.

**Table 1.** Number of cases and susceptible at non-A tables in different zones of Restaurant X. There were a total of 79 patrons on other 17 tables.

| Category of zones | Zones | Number of patrons | Number of infected cases | Attack rate (%) | *RD\**<br>(95%CI) | $X^2$ | *P* |
| --- | --- | --- | --- | --- | --- | --- | --- |
| Table A neighbours | Immediate neighbouring tables | 16 | 5 | 31.25 | 31.25<br>(8.54, 53.96) | 16.08<sup>#</sup> | <0.001<sup>#</sup> |
|  | Remote neighbouring tables | 63 | 0 | 0 |  |  |  |
| Air conditioning | ABC zone | 11 | 5 | 45.45 | 45.45<br>(16.03, 74.88) | 25.78<sup>#</sup> | <0.001<sup>#</sup> |
|  | Non-ABC zone | 68 | 0 | 0 |  |  |  |

\*RD: Rate difference

### Ventilation and dispersion of exhaled droplet nuclei

The results of two tracer gas decay experiments show that the air exchange rate was only 0.77 air changes per hour (ACH) at 16:00–17:00 and 0.56 ACH at 18:00–19:30 (Figure S4). For a volume of 431 m^3^ and 89 patrons, this is equivalent to an outdoor air supply of 1.04 and 0.75 L/s per patron, respectively.

The predicted contaminated cloud envelope in the ABC zone is shown in Figure 3. In the zone with families A, B, and C, the exhaled air stream from the index patient first falls and then rises, following the interaction of thermal plumes and the air jet of the air conditioning (Figure S2). The high-momentum air-conditioning jet carries the contaminated air at ceiling height. Upon reaching the opposite glass window, the jet bends downward and returns at a lower height. At each table, the rising thermal plumes from warm food and people carry the contaminated air upwards, and the remaining air returns to the air-conditioning unit and forms a recirculation zone or cloud envelope, referred to as the ABC zone. Similarly, other air-conditioning units also produce cloud envelopes, although these are not as distinct as that in the ABC zone, due to mixing by the air-conditioning jet of the air-conditioning unit above T09. Air exchange occurs between all of the zones because there are no physical barriers between them.

**Figure 3.**
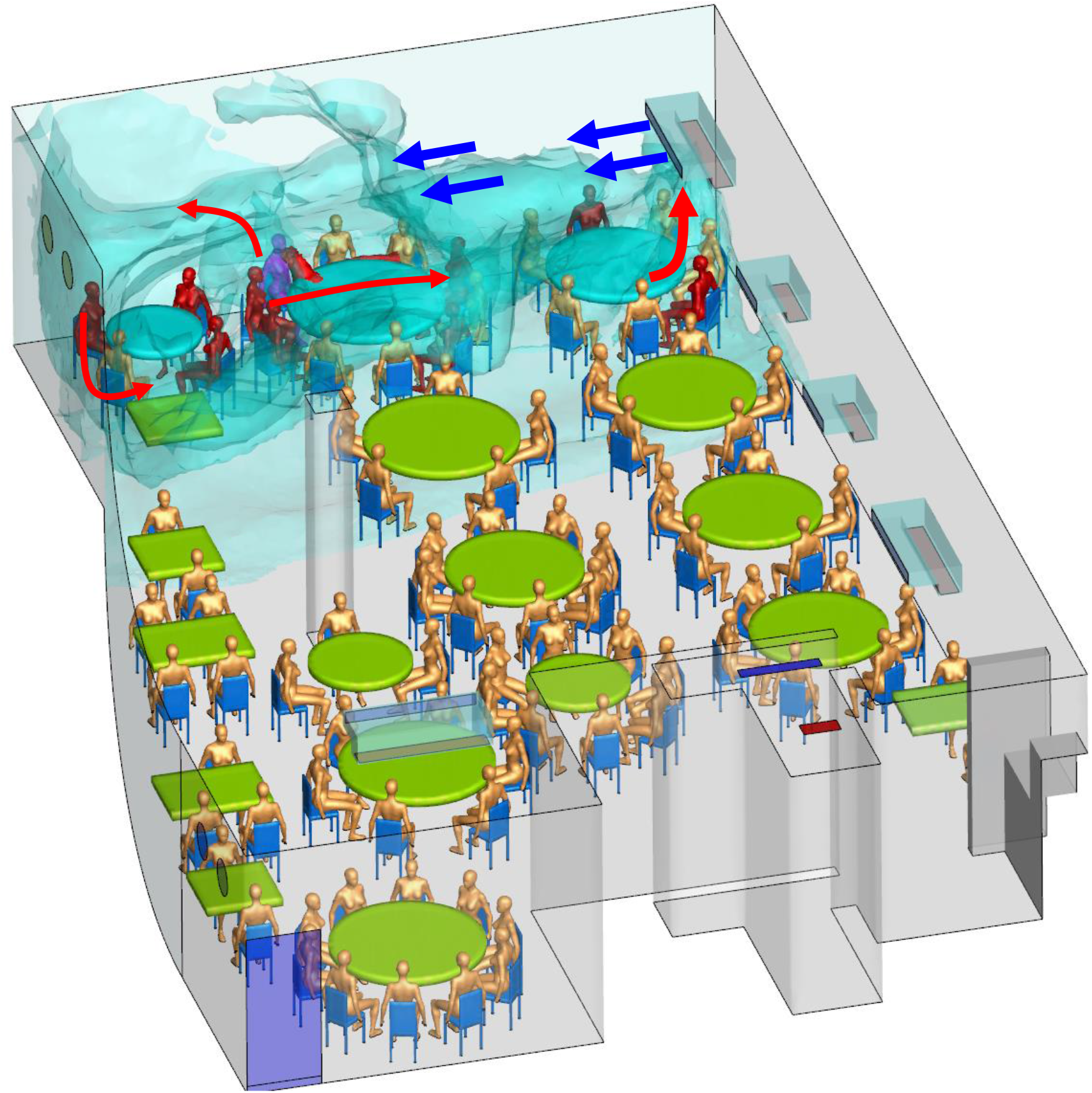
Simulated dispersion of fine droplets exhaled from index Patient A1 (magenta-blue), which are initially confined within the cloud envelope due to the zoned air-conditioning arrangement. The fine droplets eventually disperse into the other zones via air exchange and are eventually removed via the restroom exhaust fan. The ABC zone clearly has a higher concentration of fine droplets than the non-ABC zone. Other infected patients are shown in red and other non-infected in gold color. Only a single human body is used to represent all patrons

The formation of a relatively isolated contamination cloud in the ABC zone is supported by the measured ethane concentration data. The average measured ethane concentrations over a period of 66.67 min (Table S1) at TA, TB, and TC are the highest, being 1.00, 0.92, and 0.96 (normalized by concentration at TA), respectively, while the concentrations are 0.86 ppm and 0.73 ppm at T17 and T18, respectively, and are 0.55–0.70 at the other remote tables. As expected, some mixing clearly occurred between the different air-conditioning zones (Figure S1), although a stable higher concentration was maintained in the ABC zone.

The predicted average concentrations over a period of 66.67 min are listed in Table S1. According to the results of the logistic regression model, a higher measured concentration is associated with a higher risk of acquiring COVID-19 (odds ratio associated with a 1% increase in concentration: 1.115; 95% CI: 1.008–1.233; *P* = 0.035) (Table S1). Similarly, a higher predicted concentration is associated with a higher risk of acquiring COVID-19 (odds ratio associated with a 1% increase in concentration: 1.268; 95% CI: 1.029–1.563; *P* =0.026).

## Discussion

Lu et al. (2020) suggested that droplet transmission is the most likely primary cause of this outbreak, but pointed out that the outbreak cannot be explained by droplet transmission alone, because the distances between the index patient (A1) and patrons at the other tables are all greater than 1 m. We estimate that such distances may have been as far as 4.6 m (Figure S1). Lu et al. (2020) also suggested that “strong airflow from the air conditioner could have propagated droplets from table C to table A, then to table B, and then back to table C”, but stopped short of pinpointing the role of aerosol transmission due to the lack of environmental data. The role of aerosol transmission was postulated by the Chinese National Health Council (NHC) (Li and Gao, 2020) during the early phase of the COVID-19 epidemic in China, however, no specific evidence is provided in the NHC’s recommendation.

We attempted to identify the role of fomite and close contact by examining individual trajectories during the patrons’ stay in the restaurant from the available video record. However, no evidence was identified to support exposure to SARS-Co-V2 occurring via these routes in this instance.

Our prediction shows that a contaminated recirculation envelope was created in the ABC zone (Figure 3), which thus sustained a higher concentration of exhaled droplet nuclei from the index patient. The overlap period for families A and B in the restaurant was 53 min (between 12:01 and 12:54) and 75 min for families A and C (between 12:03 and 13:18), which would have allowed sufficient exposure time to the exhaled droplets. Patient C1 arrived late, at 12:32, and had a 46-min overlap with family A. That none of the waiters were infected can be attributed to their relatively short exposure time to exhaled droplets from the index patient. A relatively high concentration of trace gas was also measured at Table T17, however the patrons at this table (*n* = 5) arrived later (13:00, 18 min of exposure with Table A) and none were infected.

The formation of individual circulation zones was due to the spatial configuration and installation of five air-conditioning units (Figure S2).

However, the formation of a contaminated recirculation envelope in the ABC zone cannot alone explain the outbreak. Further evidence comes from the low ventilation rates: the observed high concentrations of the simulated contamination result from the lack of outdoor air supply. The exhaust fans in the walls were found to be turned off and sealed during the January 24 lunch, meaning that there was no outdoor air supply aside from infiltration and infrequent and brief opening of the fire door due to the negative pressure generated by the exhaust fan in the restroom. This outdoor air was mainly distributed to the non-ABC zone, thus exacerbating the ventilation deficit of the ABC zone.

The measured average air flows of 1.04 L/s and 0.75 L/s per patron in the non-ABC and ABC zones, respectively, are considerably lower than the 8–10 L/s per person required by most authorities or professional societies (American Society of Heating, Refrigerating and Air-Conditioning Engineers Standard 62.1, 2004). The restaurant was also crowded, as to accommodate the increased number of customers on Chinese New Year’s Eve, the restaurant had added extra tables. Consequently, the occupant density was only 1.55 m^2^ per patron, including the area occupied by tables. The transmission of SARS-CoV-2, which subsequently resulted in an outbreak of COVID-19, thus occurred in a crowded and poorly ventilated space.

Lack of adequate ventilation and overcrowding is known to be associated with respiratory infection outbreaks, although some are not commonly thought to be transmitted by aerosols. This restaurant SARS-CoV-2 outbreak resembles the Alaska plane influenza outbreak (Moser et al., 1979), in which a plane with a 56-seat passenger compartment was delayed by engine trouble and no mechanical ventilation was provided during the 4.5-h wait. The index patient was a passenger who became ill with influenza within 15 min after boarding the plane. There was approximately 3 m^3^ of compartment space per seat, and the provision of outdoor air was only possible by the plane doors being open for some periods during the 4.5-h wait, and during the movement of passengers in and out of the plane. According to Rudnick and Milton (2003), this resulted in there being only 0.08–0.40 L/s of air circulation per passenger, which is slightly less than the range measured in Restaurant X, and this resulted in 72% of the 54 passengers on board this plane being infected with influenza.

A systematic review by the World Health Organization (WHO) also found evidence for the association between crowding and infection (WHO, 2018). During the 2009 H1N1 pandemic, the basic reproduction number was as high as 3.0–3.6 in outbreaks in crowded schools, compared to 1.3–1.7 in less crowded settings (Lessler et al., 2009, Writing Committee, 2010). The SARS-CoV-2 virus can survive in air for at least 3 h (van Doremalen et al., 2020) and airborne influenza virus genomes and viable influenza virus particles have been detected (Lindsley et al., 2012, 2016, Yan et al., 2018, Xie et al., 2020).

It is important to note that our results do not show that long-range aerosol transmission of SARS-CoV-2 can occur in any indoor space, but that transmission may occur in a crowded and poorly ventilated space. Gao et al. (2016) showed that the relative contribution of aerosols to respiratory infection is a function of ventilation flow rate. A sufficiently high ventilation flow-rate reduces the contribution of aerosol transmission to a very low level, whereas a low ventilation flow-rate leads to a relatively high contribution of aerosols to transmission.

Both fine and large droplets are exhaled during respiration, and the infection risk from respiration is known to be highest when two people are in close contact. Liu et al. (2017) proposed that in addition to the traditional large-droplet mechanism, a short-range aerosol mechanism may also be important. An examination of the spatial concentration profile starting from the mouth of the infected person shows a continually reduced concentration profile in the exhaled jet, which weakens at some distance and merges into and becomes indistinguishable from the background room air. The average room concentration of aerosols is thus a function of source strength and ventilation rate. When the ventilation rate of the room is sufficiently low, the room average condition can become as concentrated as within the exhaled air. Hence, in theory, even if an infectious agent is not typically (i.e., under adequate ventilation) transmitted by a long-range aerosol mechanism, the spatial extent of transmission increases if the ventilation rate is very low. We refer to such transmission as an extended short-range aerosol mechanism.

In summary, our epidemiologic analysis, onsite experimental tracer measurements, and airflow simulations support the probability of an extended short-range aerosol spread of the SARS-CoV-2 having occurred in the poorly ventilated and crowded Restaurant X on January 24, 2020.

This conclusion has important implications for intervention methods in the ongoing COVID-19 pandemic. Specifically, although close contact and fomite exposure may play a major role in the transmission of SARS-CoV-2, extended short-range aerosol transmission of the virus is possible in crowded and poorly ventilated enclosures. Our study suggests that it is crucial to prevent overcrowding and provide good ventilation in buildings and transport cabins for preventing the spread of SARS-CoV-2 and the development of COVID-19.

## Data Availability

Not available

## Acknowledgements

The authors are grateful to colleagues at Guangzhou CDC, Guangzhou Yuexiu CDC, and Guangdong CDC who helped in collecting the epidemiologic data, and to Professor Bao Ruoyu and students of Sun Yat-sen University for assisting with the field experiments. The work was supported by the Research Grants Council of Hong Kong’s General Research Fund (17202719), the National Key Research and Development Program of China (2017YFC0702800), the National Natural Science Foundation of China (41875015), and the Science and Technology Planning Project of Guangdong Province (2019B111103001).

## Author contributions

Y. Li, H. Qian, J. Hang and M. Kang contributed equally. Y. Li, M. Kang, and H. Qian contributed to the study design, hypothesis formulation, and coordination. H. Qian led the CFD modeling. J. Hang led the field environmental experiments. M. Kang, J. Li, and X. Chen, contributed to the field investigation, data analyses, and reporting. X. Chen, P. Liang, and H. Ling contributed to the field studies and experiments. J. Wei and L. Liu contributed to experimental design. S. Xiao contributed to statistical analysis. Y. Li wrote the first draft of the paper. M. Kang, H. Qian, J. Hang, and X. Chen contributed major manuscript revisions. All of the other authors contributed revisions.

The authors declare no conflict of interest.

All of the authors approved the submitted version and have agreed to be personally accountable for their own contributions.

## References

1. World Health Organization (WHO) housing and health guidelines. (2018). Geneva, Switzerland: WHO, 2018. (https://www.who.int/sustainable-development/publications/housing-health-guidelines/en/.)

2. Lessler J, Reich NG, Cummings DA, New York City Department of Health and Mental Hygiene Swine Influenza Investigation Team. Outbreak of 2009 pandemic influenza A (H1N1) at a New York City school. N Engl J Med 2009; 361(27):2628–36.

3. Writing Committee of the WHO Consultation on Clinical Aspects of Pandemic (H1N1) 2009 Influenza: Clinical aspects of pandemic 2009 influenza A (H1N1) virus infection. N Engl J Med 2010; 362:1708–1719.

4. Centers for Disease Control and Prevention (CDC). How 2019-nCoV spreads. Washington DC, USA: U.S. Department of Health & Human Services, 2020. (https://www.cdc.gov/coronavirus/2019-ncov/about/transmission.html.)

5. Li X, Gao F. Public Prevention Guidelines of Infection due to the Novel Coronavirus Pneumonia (In Chinese, 新型冠状病毒感染的肺炎公众防护指南). Beijing, China: People’s Medical Publishing House, 2020:page 9.

6. Moser MR, Bender TR, Margolis HS, Noble GR, Kendal AP, Ritter DG. An outbreak of influenza aboard a commercial airliner. Am J Epidemiol 1979;110(1):1–6.

7. Rudnick SN, Milton DK. Risk of indoor airborne infection transmission estimated from carbon dioxide concentration. Indoor Air 2003;13(3):237–45.

8. Wong TW, Li CK, Tam W, Lau JTF, Yu TS, Lui SF, Chan PKS, Li YG, Bresee JS, Sung JY, Parashar UD. Cluster of SARS among medical students exposed to single patient, Hong Kong Emerg Infect Dis 2004;10:269–76.

9. Li Y, Huang X, Yu ITS, Wong TW, Qian H. Role of air distribution in SARS transmission during the largest nosocomial outbreak in Hong Kong. Indoor Air 2005;15:83–95.

10. Yu ITS, Li Y, Wong TW, Tam W, Chan A, Lee JHW, Leung DYC, Ho T. Evidence of airborne transmission of the severe acute respiratory syndrome virus. N Engl J Med 2004;350:1731–9.

11. Lu J, Gu J, Li K, et al. COVID-19 outbreak associated with air conditioning in restaurant, Guangzhou, China. Emerg Infect Dis 2020;26(7). https://doi.org/10.3201/eid2607.200764

12. van Doremalen N, Morris DH, Holbrook MG, et al. Aerosol and surface stability of SARS-CoV-2 as compared with SARS-CoV-1. N Engl J Med 2020 doi:10.1056/NEJMc2004973.

13. Yan J, Grantham M, Pantelic J, et al. Infectious virus in exhaled breath of symptomatic seasonal influenza cases from a college community. P Natl Acad Sci USA 2018;115:1081–6.

14. Lindsley WG, Blachere FM, Beezhold DH, et al. Viable influenza A virus in airborne particles expelled during coughs versus exhalations. Influenza Other Resp 2016;10:404–13.

15. Lindsley WG, Pearce TA, Hudnall JB, et al. Quantity and size distribution of cough-generated aerosol particles produced by influenza patients during and after illness. J Occup Environ Hyg 2012;9(7):443–9.

16. Xie C, Lau EH, Yoshida T, et al. Detection of influenza and other respiratory viruses in air sampled from a university campus: a longitudinal study. Clin Infect Dis 2020;70(5):850–8.

17. American Society of Heating, Refrigerating and Air-Conditioning Engineers (ASHRAE) Ventilation for acceptable indoor air quality. Atlanta, USA: ASHRAE Standard 62.1, 2004.

18. Bivolarova M, Ondráček J, Melikov A & Ždímal V. A comparison between tracer gas and aerosol particles distribution indoors: The impact of ventilation rate, interaction of airflows, and presence of objects. Indoor Air 2017; 27(6):1201–1212.

